# A plasma metabolite score of three eicosanoids predicts incident type 2 diabetes – a prospective study in three independent cohorts

**DOI:** 10.1101/2021.07.23.21260986

**Authors:** Karolina Tuomisto, Joonatan Palmu, Tao Long, Jeramie D. Watrous, Kysha Mercader, Kim A. Lagerborg, Allen Andres, Marko Salmi, Sirpa Jalkanen, Ramachandran S. Vasan, Michael Inouye, Aki S. Havulinna, Pekka Jousilahti, Teemu J. Niiranen, Susan Cheng, Mohit Jain, Veikko Salomaa

## Abstract

**Aim:** Peptide markers of inflammation have been associated with the development of type 2 diabetes. The role of upstream, lipid-derived mediators of inflammation such as eicosanoids, remains less clear. The aim was to examine whether eicosanoids are associated with incident type 2 diabetes.

**Methods:** In the FINRISK 2002 study, a population-based sample of Finnish men and women aged 25-74 years, we used directed, non-targeted liquid chromatography – mass spectrometry to identify 545 eicosanoids and related oxylipins in the participants’ plasma samples (n=8,292). We used multivariable-adjusted Cox regression to examine associations between eicosanoids and incident type 2 diabetes. The findings were replicated in the Framingham Heart Study (FHS, n=2,886) and DILGOM 2007 (n=3,905). Together, these three cohorts had 1070 cases of incident type 2 diabetes.

**Results:** 76 eicosanoids were associated individually with incident type 2 diabetes. We identified three eicosanoids independently associated with incident type 2 diabetes using stepwise Cox regression with forward selection and a Bonferroni-corrected inclusion threshold. A three-eicosanoid risk score produced a hazard ratio (HR) of 1.56 (95% confidence interval 1.41-1.72) per one standard deviation (SD) increment for risk of incident diabetes. The HR for comparing the top quartile to the lowest was 2.80 (2.53-3.07). Meta-analysis of the three cohorts yielded a pooled HR per SD of 1.31 (1.05-1.56).

**Conclusion:** Plasma eicosanoid profiles predict incident type 2 diabetes and the clearest signals replicate in three independent cohorts. Our findings give new information on the biology underlying type 2 diabetes and suggest opportunities for early identification of people at risk.

## INTRODUCTION

Diabetes and its complications contribute significantly to the global burden of disease: in 2017, it was the third leading cause of global Years Lost due to Disability for men and the fifth for women [1]. It is estimated that its global costs (1.3 trillion US$ in 2015) will continue to rise by 2030, even if Sustainable Development Goals related to the disease are met [2].

The prevention of complications, such as cardiovascular disease, diabetic nephropathy, neuropathy and retinopathy, lie at the heart of diabetes care. However, clinical trials have shown that the onset of type 2 diabetes can be postponed, and even prevented, by lifestyle changes, such as changes in the intake of fats and fibers, and increasing physical activity, and medical treatment, such as metformin medication [3,4]. Therefore, better understanding of factors involved in the development of type 2 diabetes, together with earlier identification of people at risk, are important for optimal targeting of right preventive measures to right people.

Type 2 diabetes is a complex disease involving interactions of both environmental risk factors and genetics, and, at an undetermined scale, of inflammation. Type 2 diabetes is closely linked to cardiovascular and kidney diseases, obesity and the metabolic syndrome, all of which are known to be associated with low-grade inflammation. Biomarkers of systemic inflammation, such as cytokines, adiponectin and high-sensitivity C-reactive protein (hs-CRP), have been associated with the development of type 2 diabetes in large population-based studies and randomized controlled trials [5,6,7,8,9].

Situated proximally (upstream) in the inflammation pathways is a large group of bioactive lipids known as eicosanoids and related oxylipins. These chemical factors are derived from arachidonic acid and other polyunsaturated fatty acids (PUFAs) by enzymatic mechanisms, notably via cyclooxygenase, lipoxygenase and cytochrome P450 enzymes, as well as by non-enzymatic mechanisms. Eicosanoids mediate inflammatory processes directly at local level as well as systemically, including through modulation of cytokines and other inflammatory mediators [10]. With recent advances in mass spectrometry-based metabolomics, hundreds of eicosanoid and oxylipin species can now be detected and quantified at a rapid scale in human plasma [11,12].

The aim of this study was to examine whether plasma eicosanoid profiles are associated with risk of incident type 2 diabetes in a prospective follow-up of a population-based Finnish cohort. The significant findings were replicated in an independent cohort from the North American sub-continent and in another population-based Finnish cohort. Together, these cohorts included 1070 cases of incident type 2 diabetes. A secondary aim was to investigate whether the eicosanoid effect is mediated by inflammation or insulin resistance.

## MATERIAL AND METHODS

### Population, data collection and design

#### FINRISK 2002

FINRISK 2002 study is a population-based random sample of individuals aged 25-74 years living in Finland (n=8798, participation rate 65.2%). Sampling included stratification by sex, region and 10-year age groups. Participants responded to questionnaires, underwent physical examination by trained nurses and gave blood samples. Detailed study protocols, including the sampling, measurements and blood sample protocols have been previously described [13]. Plasma samples from 8,292 participants were successfully analyzed with directed non-targeted Liquid Chromatography - Mass Spectrometry (LC-MS), as described in previous publications [11,12]. In addition, an oral glucose tolerance test (OGTT) was carried out in 3,767 participants (45 years of age and older) and data on glucose tolerance status was obtained for 3,092 participants without prior history of diabetes. A panel of cytokines was quantified from blood samples of 2,951 participants.

For the FINRISK 2002 cohort, prevalent diabetes cases at baseline and incident cases during follow-up were identified combining information from the National Hospital Discharge Register (NHDR), Causes of Death Register (CDR) and the Drug Reimbursement and Drug Purchase Registers (DPR), using the Finnish personal identification number, up to December 31^st^, 2017. Diagnostic codes for extracting information from NHDR and CDR were E10-E14 (ICD10) or 250*B (ICD9). Diabetes medication purchase ATC code A10 or a special reimbursement code for diabetes medications were used for identifying diabetes cases from the Drug Purchase register. If medicine purchase was the only fulfilled criterion, ≥3 purchases were required and diagnosis codes were used to exclude gestational diabetes. To determine the type of diabetes, we used a proxy variable: all participants under 30 years of age and treated with insulin only, or in combination with metformin, and those aged 30-40 years when insulin only was started, were categorized as type 1 diabetes. All other persons with diabetes were considered to have type 2 diabetes. Thanks to the country-wide electronic health care registers, the follow-up was virtually 100% complete.

Participants with prevalent or incident type 1 diabetes were excluded from the analysis (n=79). Record linkages based on the personal identification code to NHDR, CDR and DPR were also used to identify subjects to be excluded from the analysis, due to cancer (excluding ICD10 category C44, n=1,085). Participants pregnant at baseline (n=44) were also excluded from the final analyses. Finally, after excluding further participants with missing values of variables relevant for our analyses, the total study population consisted of 6,548 participants. Supplement Fig S1 presents a flowchart formulating the final study sample.

#### FHS Offspring

FHS Offspring study is a cohort of 5,124 participants that were first examined in 1971 and consecutively re-examined every four to eight years. The cohort consists of the children of the FHS first-generation cohort, and their spouses. Participants of the original cohort were enrolled in a longitudinal community-based cohort study in 1948, including a random sample of two thirds of the adult population of Framingham, MA. Detailed study protocols including sampling have been previously published [14]. For this study, we considered individuals who participated in the eighth examination cycle of FHS Offspring in 2005– 2008 and whose samples were successfully analyzed with LC-MS (n=2,886). After excluding 771 participants with diabetes at baseline or missing covariates, we included n=2,115 participants as the replication cohort. Diabetes mellitus in FHS Offspring cohort was diagnosed either by fasting plasma glucose ≥126 mg/dL (7.0 mmol/l), non-fasting plasma glucose ≥200 mg/dL (11.0 mmol/l), or treatment with insulin or an oral hypoglycemic agent as ascertained at routine FHS examinations or based on annual medical health history updates.

#### DILGOM 2007

The DILGOM (the DIetary, Lifestyle and Genetic determinants of Obesity and Metabolic syndrome) study is an extension of the FINRISK 2007 survey, which included a random population-based sample of individuals aged 25-75 years living in Finland. All participants of the FINRISK 2007 survey were invited to take part in the more focused DILGOM study and 5024 participated (participation rate 80%).

Participants responded to self-administered questionnaires, underwent a physical examination by trained nurses and gave blood samples. Detailed study protocols, including sampling, measurements and blood sample protocols have been previously described [15]. Plasma samples from n=4,903 participants were successfully analyzed with LC-MS. Cases of diabetes were identified and information from registries combined in an identical way to the FINRISK 2002 cohort, as described above. Similarly, participants with prevalent type 1 diabetes, pregnancy, cancers (excluding ICD10-C44) and missing values were excluded from the final replication cohort. Finally, 3,905 individuals were included in the analyses.

### Ethics statement

The FINRISK 2002 study was approved by the Ethical Committee for Epidemiology and Public Health of the Helsinki University Hospital District on December 19, 2001. The DILGOM 2007 study was approved by the Coordinating Ethical Committee of the Helsinki and Uusimaa Hospital District on April 03, 2007. FHS Offspring study was approved by Boston University Medical Center’s Institutional Review Board.

### Laboratory methods and plasma eicosanoid profiling

Plasma eicosanoid profiling for all three cohorts was carried out in the same laboratory at the University of California San Diego, USA. The profiling methodology is described in more detail elsewhere [11,12]. In short, we used directed, non-targeted LC-MS combined with computational chemical networking to identify eicosanoids in participants’ plasma samples. In the discovery sample, FINRISK 2002, a total of 545 eicosanoids and related oxylipins were validated using methodologies such as spectral fragmentation pattern networking and manual annotation.

The OGTT was carried out according to the World Health Organization (WHO) recommendations, and the testing and measurement methodologies have previously been described in detail [16]. Fasting plasma glucose and insulin concentrations were used to calculate the homeostasis model assessment for insulin resistance (HOMA-IR) and for beta cell function (HOMA-B) indices [17].

We quantified well-known inflammatory markers often hypothesized to associate with diabetes. High-sensitivity C-reactive protein (hs-CRP) was quantified with the Architect ci8200 Chemistry Analyser (Abbott Laboratories, USA) from FINRISK 2002 serum samples and with Architect c8000 analyzer (Abbott Laboratories, USA) from DILGOM 2007 serums samples, using an immunoturbidometric method (Sentinel diagnostics, Italy). Interleukin-1 receptor antagonist (IL-1Ra), interleukin-6 (IL-6) and tumor necrosis factor alpha (TNF-alpha) were quantified from previously unthawed heparin plasma samples using Bio-Rad’s premixed Bio-Plex Pro Human Cytokine 27-plex Assay and 21-plex Assay, and Bio-Plex 200 reader with Bio-Plex V.6.0 software (Bio-Rad Laboratories, USA). The detailed methodology has been previously published [18,19].

### Statistical methods

The eicosanoid profiling data was normalized using plate medians corrected for plate deviation: plate medians were subtracted from each feature and then divided by the median absolute deviation [20].

We used means, or where relevant due to skewed distributions, geometrical means, and interquartile ranges to summarize baseline characteristics of continuous variables and frequencies for categorical variables. For subsequent analyses, participants with prevalent type 2 diabetes (n=167) were excluded from the FINRISK study population resulting in n=6,381 (with n=586 of incident type 2 diabetes cases).

To test the association of eicosanoids with incident type 2 diabetes, we used Cox proportional hazards regression and a nested modeling approach, adjusting for well-established risk factors for type 2 diabetes, and other confounding factors, in three different models. The first model adjusted for age, sex, region of residence, and mass spectrometry plate. The second model (model 2) added adjustments for BMI, physical activity, parental history of diabetes, prevalent CVD, systolic blood pressure, antihypertensive medication, triglycerides and lipid lowering medication. We also ran a third model that was further adjusted for hs-CRP. The proportional hazards assumption was tested using Schoenfeld residuals [21]. We used the False Discovery Rate (FDR) correction method for type I error control in multiple comparisons [22]. We also produced a correlation heatmap for eicosanoids significantly associated with incident type 2 diabetes, ordered using the complete linkage clustering method. Correlations with hs-CRP were also added to the heatmap.

Eicosanoids with significant associations in any of the three longitudinal analysis models (described above) were then included in a stepwise Cox regression analysis with forward selection, applying a Bonferroni-corrected inclusion threshold of p<0.05/545 = 0.00009 [23]. We constructed an eicosanoid risk score for the three remaining eicosanoids using their respective regression coefficients as weights in the prediction model.

We examined type 2 diabetes-free survival across quartiles of the risk score using Cox models and Kaplan-Meier survival curves. We compared survival distributions using the log rank test [24]. Using Cox regression, we calculated the risk per standard deviation (SD) change in the eicosanoid risk score for the different models.

We calculated medians of fP-Glucose (fP-Gluc), fP-Insulin, HOMA-IR and HOMA-B for the eicosanoid score quartiles and tested the linear trends across the quartiles adjusting for age, sex and BMI. We explored the correlations between fP-Gluc, fP-Insulin, HOMA-IR, HOMA-B and three eicosanoids from the risk score using Spearman correlation. Correlations were also calculated between a panel of cytokines, namely IL-1Ra, IL-6, TNF-alpha and the same three eicosanoids.

Replication analyses for the three eicosanoids exceeding the Bonferroni-corrected inclusion threshold (p<0.05/545 = 0.00009) in FINRISK were performed in the FHS Offspring cohort with Cox regression adjusted for age, sex, BMI, systolic blood pressure, hypertension treatment, and triglycerides. Participants with baseline diabetes (n=302) and missing values in the covariates were excluded from the analysis, resulting in n=236 incident diabetes cases out of a total of 2,115 individuals included.

In the DILGOM 2007 cohort, we excluded participants with prevalent type 2 diabetes (n=144), and thus 3,761 individuals (out of which n=248 with incident type 2 diabetes) were included in the replication analysis. The analysis was performed with Cox regression adjusting for the same covariates as in the FINRISK cohort.

Meta-analysis of the risk scores in the three cohorts was performed using a Random-Effects Model (*metafor* package in R). We used R, version 3.6.1, for all analyses and the source code for analyses is available at https://doi.org/10.5281/zenodo.3968712.

## RESULTS

Baseline characteristics of the FINRISK 2002, DILGOM 2007 and FHS Offspring cohort participants (women and men) are presented in Table 1. HbA1c values were only available for FINRISK participants older than 50 years (n=3,586).

**Table 1.** Baseline characteristics of FINRISK 2002 discovery cohort (n=6, 548) and FHS Offspring (n=2, 886) and DILGOM 2, 007 (n=3, 905) replication cohorts.

|  | FINRISK 2002 |  | Replication cohort (FHS) |  | Replication cohort (DILGOM) |  |
| --- | --- | --- | --- | --- | --- | --- |
|  | Women<br>n=3,423 | Men<br>n=3,125 | Women<br>n=1,571 | Men<br>n=1,315 | Women<br>n=2,059 | Men<br>n=1,846 |
| Age, years | 45.6 (20.8) | 46.6 (19.9) | 66.4 (9.0) | 66.3 (9.0) | 49.8 (13.4) | 50.7 (13.1) |
| Weight, kg | 69.5 (16.6) | 84.5 (17.3) | 71.5 (15.8) | 88.5 (15.9) | 70.7 (13.9) | 84.7 (13.9) |
| Height, m | 1.63 (0.08) | 1.76 (0.09) | 1.61 (0.06) | 1.75 (0.07) | 1.63 (0.06) | 1.76 (0.07) |
| BMI, kg/m<sup>2</sup> | 26.3 (6.2) | 27.2 (5.1) | 27.7 (5.9) | 29.0 (4.7) | 26.7 (5.3) | 27.3 (4.1) |
| Waist-to-hip ratio | 0.84 (0.08) | 0.96 (0.09) |  |  | 0.86 (0.06) | 0.97 (0.07) |
| Mean systolic blood pressure, mmHg | 131 (26) | 136 (22) | 128 (18) | 129 (17) | 133 (21) | 138 (19) |
| HbA1c, mmol/mol | 36.2 (5.0)<br>n=1,840 | 36.4 (5.0)<br>n=1,758 |  |  |  |  |
| HDL, mmol/l <sup>a</sup> | 1.59 (0.53) | 1.30 (0.43) | 1.65 (0.47) | 1.28 (0.37) | 1.53 (0.49) | 1.27 (0.40) |
| LDL, mmol/l <sup>a</sup> | 3.07 (1.11) | 3.38 (1.20) | 2.84 (0.82) | 2.58 (0.78) | 3.04 (1.13) | 3.12 (1.16) |
| Triglycerides, mmol/l <sup>a</sup> | 1.07 (0.63) | 1.41 (1.01) | 1.31 (0.69) | 1.36 (0.88) | 1.08 (0.66) | 1.40 (0.98) |
| hs-CRP, mmol/l <sup>a</sup> | 1.15 (2.07) | 1.10 (1.66) |  |  | 1.17 (2.00) | 1.09 (1.63) |
| Current smokers, n (%) | 761 (22.3%) | 1006 (32.3%) | 147 (9.4%) | 111 (8.5%) | 307 (14.9%) | 411 (22.2%) |
| **Education, n (%)** |  |  |  |  |  |  |
| Low | 1,132 (33.6%) | 1,071 (34.8%) |  |  | 656 (31.9%) | 522 (28.3%) |
| Middle | 1,105 (32.8%) | 1,001 (32.5%) |  |  | 689 (33.5%) | 646 (35.0%) |
| High | 1,131 (33.6%) | 1,004 (32.6%) |  |  | 693 (33.7%) | 668 (36.2%) |
| **Physical activity at leisure time, n (%)** |  |  |  |  |  |  |
| Low level or no exercise | 777 (22.7%) | 718 (23.0%) |  |  | 374 (18.2%) | 345 (18.7%) |
| Light exercise, at least 4h per week | 1893 (55.3%) | 1585 (50.7%) |  |  | 1111 (54.0%) | 936 (50.7%) |
| Aerobic exercise, at least 3h per week, or regular exercise at competitive level | 753 (22.0%) | 822 (26.3%) |  |  | 574 (27.9%) | 565 (30.6%) |
| Use of antihypertensive medicines, n (%) | 428 (12.5%) | 376 (12.0%) | 699 (44.6%) | 701 (53.6%) | 357 (17.3%) | 341 (18.5%) |
| Use of lipid lowering medication, n (%) | 220 (6.4%) | 224 (7.2%) |  |  | 229 (11.1%) | 298 (16.1%) |
| Prevalent CVD, n (%) | 48 (1.4%) | 126 (4.0%) |  |  | 38 (1.8%) | 94 (5.1%) |
| Family history of diabetes, n (%) | 897 (26.2%) | 731 (23.4%) |  |  | 629 (30.5%) | 474 (25.7%) |
| Prevalent T2DM, n (%) | 80 (2.3%) | 87 (2.8%) | 135 (8.6%) | 166 (12.6%) | 52 (2.5%) | 92 (4.9%) |

Means (interquartile ranges) for continuous variables, frequencies (percentage) for categorical variables; <sup>a</sup>geometric means used due to skewed distribution

### Longitudinal analyses: independent eicosanoids

For incident type 2 diabetes, we observed 76 significant associations using the adjustment models described in the methods section. Eicosanoids, their FDR-corrected levels of significance and the direction of the association with type 2 diabetes, are visualized in Fig 1. The heatmap of pairwise correlations for the 76 eicosanoids with significant associations, and hs-CRP, is shown in Supplement Fig S2. Overall, the eicosanoids correlated with each other, with some clusters of strong positive correlations.

**Figure 1.**
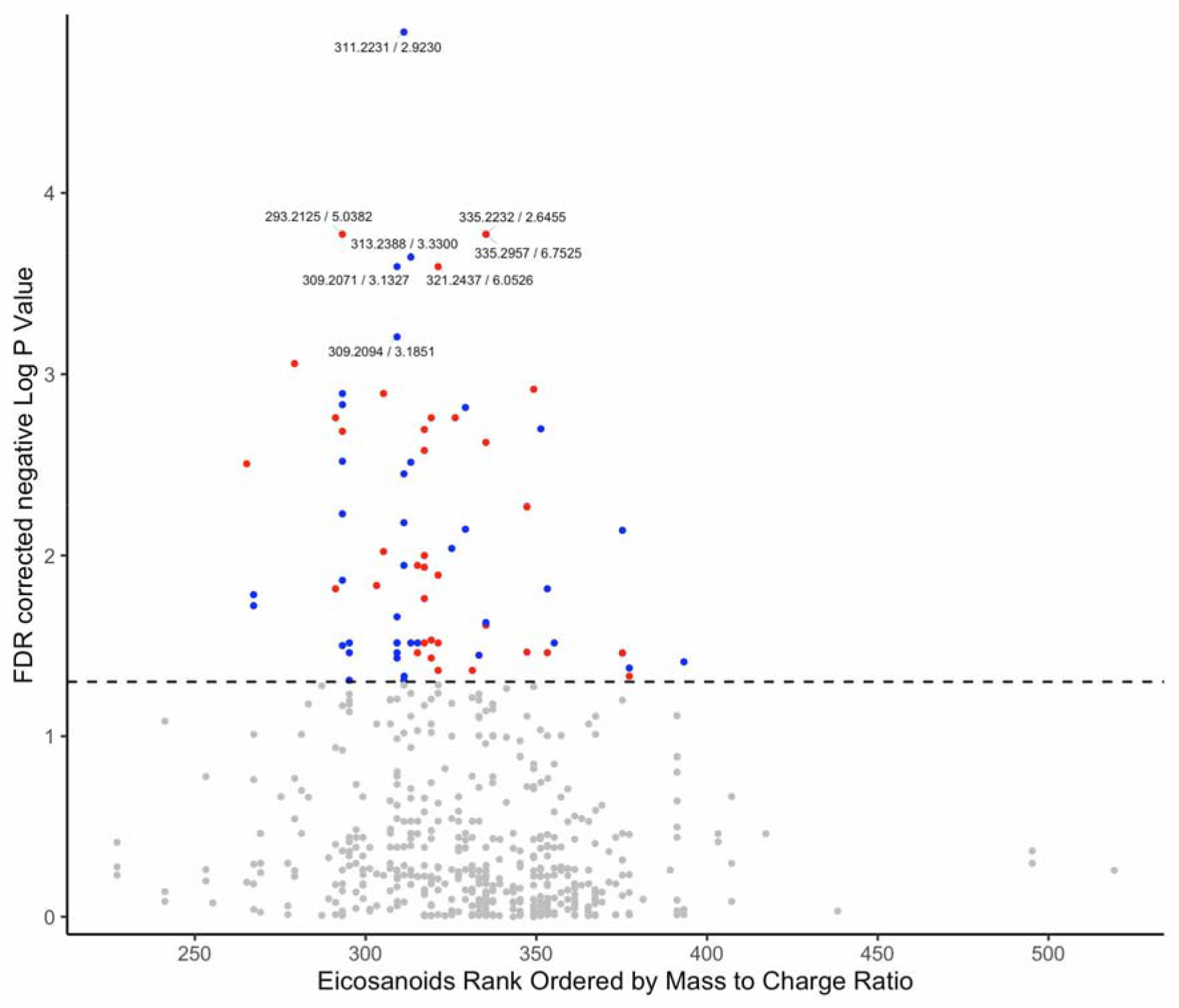
Associations between eicosanoids and incident type 2 diabetes in FINRISK 2002. A statistically significant association was found for 76 eicosanoids and incident type 2 diabetes: red is for positive associations, blue is for negative associations, grey for nonsignificant. Dashed line represents the FDR corrected level of significance. Analyses adjusted for age, sex, BMI, physical activity, parental history of diabetes, prevalent CVD, systolic blood pressure, antihypertensive medication, triglycerides, lipid lowering medication, geographical area and mass spectrometry plate

We included 76 eicosanoids that showed significant associations with incident type 2 diabetes in the stepwise Cox regression analysis using model 2. Three eicosanoids remained in the model, notably an unknown eicosanoid (EIC 62), 8-iso-Prostaglandin A1 (8-iso-PGA1) and 12-Hydroxy-5,8,10-heptadecatrienoic acid (12-HHTrE). The forest plot (Fig 2) presents hazard ratios (HR) and 95% confidence intervals (95%CI) for the individual eicosanoids remaining in the model. The unknown eicosanoid was inversely associated with incident type 2 diabetes, whereas 8-iso-PGA1 and 12-HHTrE were positively associated with the disease risk.

**Figure 2.**
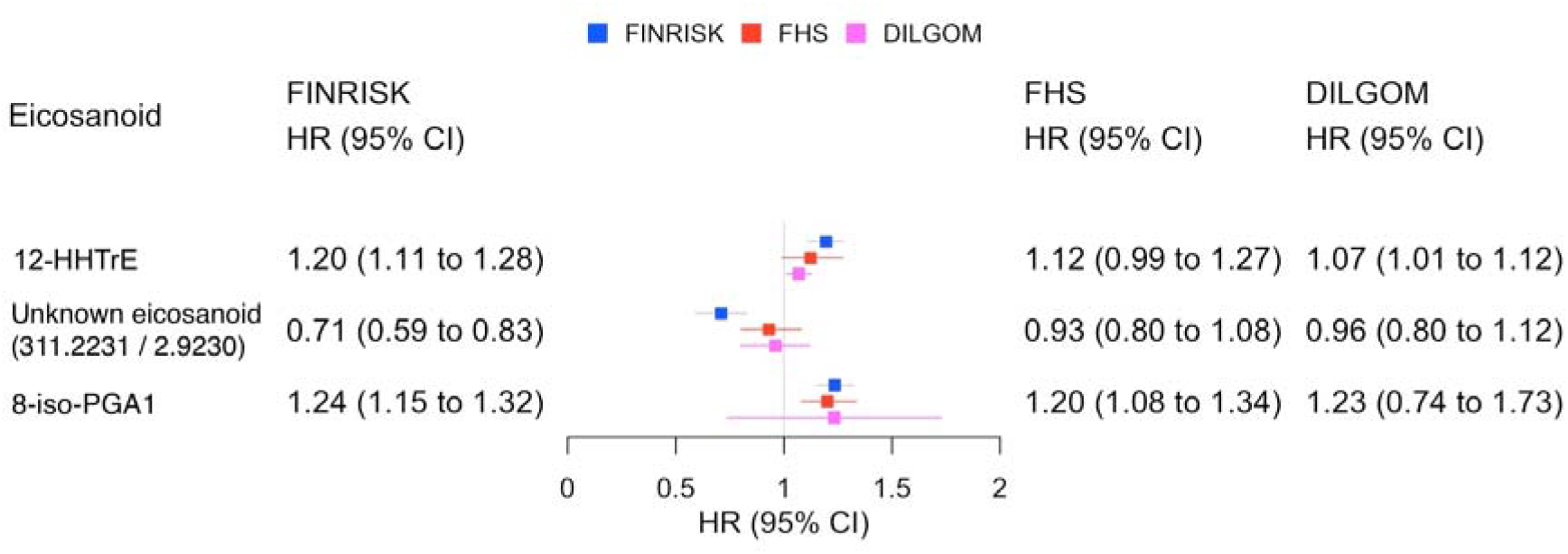
Forest plot showing hazard ratios and 95% confidence intervals for three eicosanoids independently associated with incident type 2 diabetes. Cox proportional hazards regression analyses for the FINRISK and DILGOM cohorts are adjusted for age, sex, BMI, physical activity, parental history of diabetes, prevalent CVD, systolic blood pressure, antihypertensive medication, triglycerides, lipid lowering medication, geographical area and mass spectrometry plate. For the FHS cohort, Cox proportional hazards regression analyses are adjusted for age, sex, BMI, systolic blood pressure, hypertension treatment, and triglycerides

Cox regression on the three eicosanoids was performed in the replication cohorts and the results are shown in Fig 2. The eicosanoid 8-iso-PGA1 was replicated in the FHS (p<0.001) and in the DILGOM (p=0.025) cohorts, and the other two were directionally consistent.

### Longitudinal analyses: eicosanoid risk score

Using these three eicosanoids, we established an eicosanoid risk score. Results are presented for women and men combined as testing revealed no interaction with sex and the eicosanoid risk score. The multivariate adjusted hazard ratio per SD for the three different cohorts are presented in Fig 3, together with the results of a random effects meta-analysis of the risk scores in the three cohorts.

**Figure 3.**
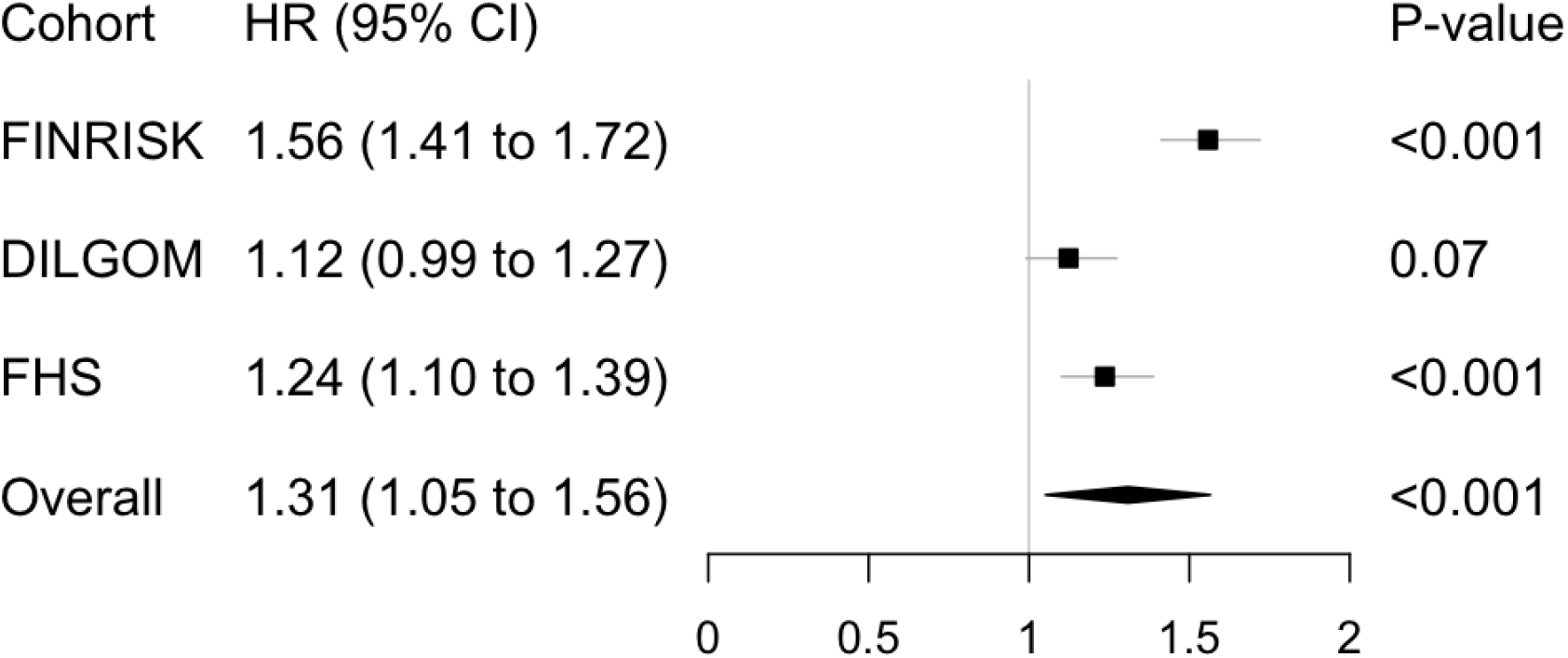
Forest plot showing hazard ratios and 95% confidence intervals for the eicosanoid risk score and its independent association with incident type 2 diabetes in the FINRISK cohort, FHS and DILGOM replication cohorts and their meta-analysis. Cox proportional hazards regression analyses for the FINRISK and DILGOM cohorts are adjusted for age, sex, BMI, physical activity, parental history of diabetes, prevalent CVD, systolic blood pressure, antihypertensive medication, triglycerides, lipid lowering medication, geographical area and mass spectrometry plate. For the FHS cohort, Cox proportional hazards regression analyses are adjusted for age, sex, BMI, systolic blood pressure, hypertension treatment, and triglycerides. Meta-analysis was calculated using the Random-Effects Model.

Hazard ratios for risk quartiles are presented in Table 2. In the top risk quartile of participants, compared to the lowest quartile of the risk score, the hazard ratio was 2.80 (95%CI 2.53-3.07) in FINRISK, 1.74 (1.31-2.31) in FHS and 1.37 (95%CI 1.00-1.74) in DILGOM. The respective HRs for quartiles 2 and 3 demonstrated a dose-response type relationship of the eicosanoid score with the risk of incident type 2 diabetes in all three cohorts. Furthermore, Kaplan-Meier curves for the risk score quartiles in FINRISK are presented in Fig 4. Disease-free survival in the lowest quartile was 95.7% and 82.6% in the top quartile.

**Table 2.** Hazard ratios and 95% confidence intervals by quartile of the eicosanoid risk score for incident type 2 diabetes in FINRISK 2002, FHS Offspring and DILGOM 2007.

| Risk quartile (Q) | FINRISK 2002 |  |  | FHS Offspring |  |  | DILGOM 2007 |  |  |
| --- | --- | --- | --- | --- | --- | --- | --- | --- | --- |
|  | HR | 95% CI | p-value | HR | 95% CI | p-value | HR | 95% CI | p-value |
| **Q1** | 1.00 | Reference category |  | 1.00 | Reference category |  | 1.00 | Reference category |  |
| **Q2** | 1.31 | 0.99-1.63 | 0.098 | 1.11 | 0.81-1.53 | 0.500 | 1.14 | 0.75-1.54 | 0.512 |
| **Q3** | 1.88 | 1.59-2.17 | <0.0001 | 1.38 | 1.03-1.85 | 0.030 | 1.30 | 0.93-1.67 | 0.165 |
| **Q4** | 2.80 | 2.53-3.07 | <0.0001 | 1.74 | 1.31-2.31 | 0.0001 | 1.37 | 1.00-1.74 | 0.093 |

**Figure 4.**
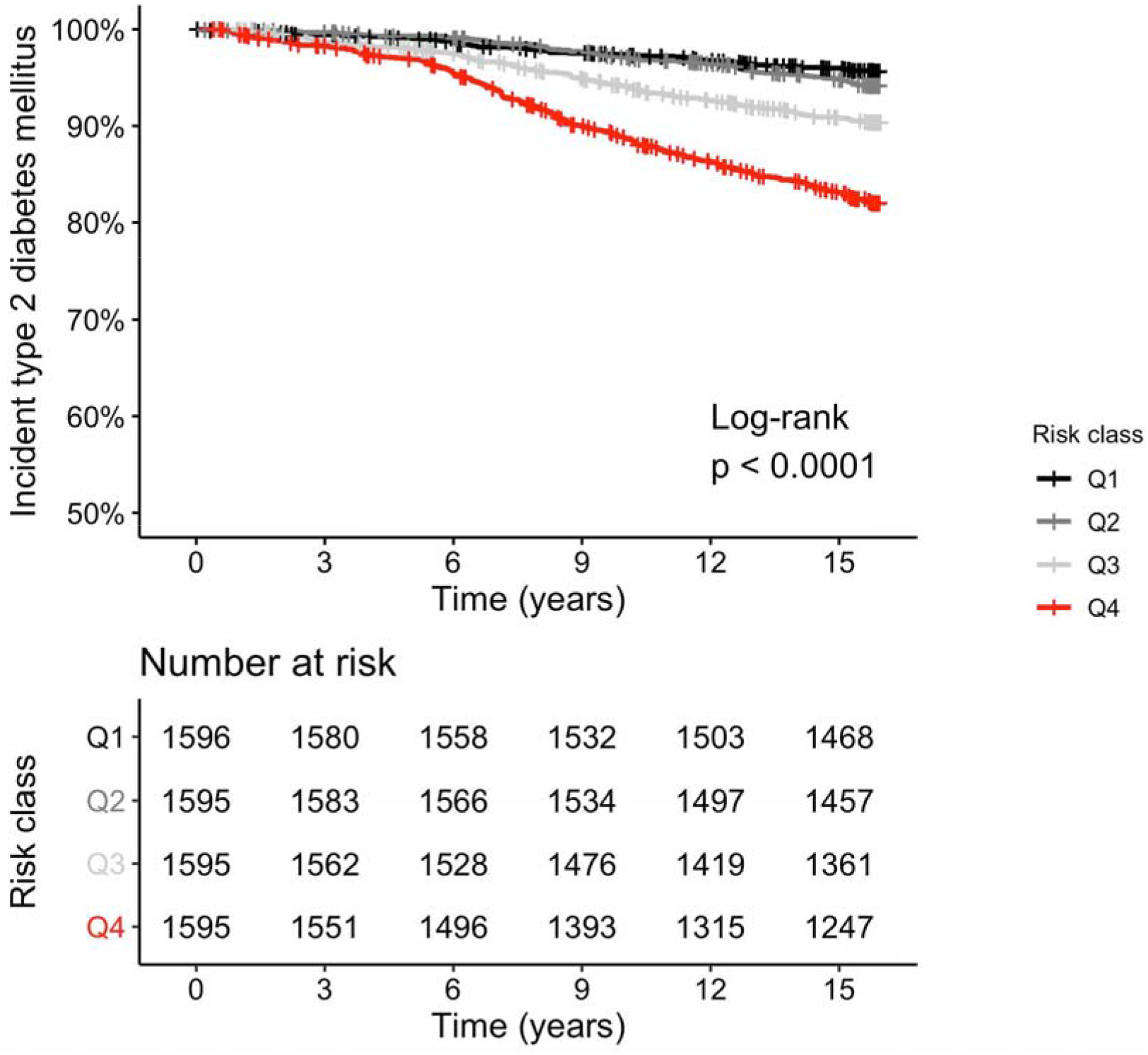
Kaplan-Meier curves for the three-eicosanoid risk score and incident type 2 diabetes in FINRISK 2002. Diabetes-free survival analysis for the different risk classes/quartiles (Q1-Q4). Analyses adjusted for age, sex, BMI, physical activity, parental history of diabetes, prevalent CVD, systolic blood pressure, antihypertensive medication, triglycerides, lipid lowering medication, geographical area and mass spectrometry plate

### Eicosanoid associations with glucose and insulin levels and commonly known cytokines

The medians of fP-Gluc, fP-Ins, HbA1c and HOMA-IR increased across the eicosanoid risk score quartiles, and the increasing trend remained statistically significant (p<0.0001) after adjusting the analyses for age, sex and BMI. For HOMA-B, the increase was less significant (p=0.014). (Supplement Table S1).

Correlations of the three eicosanoids with a panel of cytokines are presented in Supplement Table S2. The eicosanoid identified as 12-HHTrE was quite strongly correlated with fP-Gluc, fP-Ins, HbA1c, HOMA-IR, HOMA-B and hs-CRP. The other eicosanoids showed some statistically significant correlations with these glucose metabolism indicators but the magnitudes of the correlation coefficients were modest.

None of the three eicosanoids correlated with the blood concentrations of inflammatory cytokines. In addition, Cox proportional hazards regression analysis for the multivariable model, further adjusted for the inflammatory cytokines, remained significant for both the continuous three-eicosanoid risk score as well as the 3rd and 4th risk quartiles of the score.

## DISCUSSION

Downstream mediators of inflammation such as CRP and other peptide mediators of inflammation such as cytokines, have received a lot of attention in the study of the development of type 2 diabetes [25]. Previous studies have pointed to adiponectin, apolipoprotein B, CRP, IL1-Ra and ferritin playing a role in type 2 diabetes prediction [6,7]. The Whitehall II study examining MR/CRP haplotypes suggested non-causal associations between CRP, insulin resistance, glycemia and diabetes, and that upstream effectors of inflammation may play a more causal role in the development of diabetes [26].

Combined with advanced metabolomics methodologies, our statistically cogent survival analysis using a three-eicosanoid risk score suggests a significant independent role for lipid-derived, upstream mediators of inflammation in the prediction of incident type 2 diabetes. One of the three eicosanoids showed an independent inverse association with incident type 2 diabetes, suggesting potential protective characteristics for some plasma eicosanoids. However, this negative association was not statistically significant in the replication cohorts.

In 2011, Luo & Wang examined the role of eicosanoids from the different arachidonic acid metabolism pathways in pathogenesis of diabetes: they proposed that eicosanoids metabolized by the cyclooxygenase (COX) pathway are active participants in the function of beta-cells, those metabolized by the lipoxygenase (LOX) pathway act as mediators in the beta-cell inflammation whereas the role of eicosanoids metabolized via the cytochrome P450 enzymes was still unclear [27]. More recently, metabolites have been suggested to act as regulators of insulin sensitivity and further hypothesized to play a role in the pathogenesis of insulin resistance and type 2 diabetes [28].

Out of the individual eicosanoids identified for our three-eicosanoid risk score, two are metabolites with known bioactivity. 8-iso-PGA1 is part of isoprostanes, which are stereoisomers of prostaglandins formed independent of COX enzymes, through peroxidation of arachidonic acid. They are considered as markers of oxidative stress, which may lead to chronic inflammation [29,30]. Studies have suggested a causal role for isoprostanes in the development of different disease states, such as incident type 2 diabetes [30,31].

Oxidative stress has also been implicated in the development of complications of diabetes such as nephropathy and neuropathy [32,33]. A recent study found that individuals with heredity of type 2 diabetes have an increased susceptibility to oxidative stress and inflammation following a high-carbohydrate meal [34].

The second known eicosanoid demonstrating a positive independent association with incident type 2 diabetes is 12-HHTrE. Different enantiomers of 12-HHTrE are metabolites of nonenzymatic activity as well as of COX- and LOX-pathways, and they have been implicated in conditions such as breast cancer, hypertension, endothelial prostacyclin formation, response to acute exercise and high carbohydrate intake [35,36,37,38].

A study using the same mass spectrometry platform recently found 12-HHTrE to be robustly associated with blood pressure [36]. None of the eicosanoids associated with blood pressure were correlated with 8-iso-PGA1, however one of them (Tetranor-12(R)-HETE) strongly correlated with 12-HHTrE. Furthermore, the unknown eicosanoid from our risk score showed strong correlation with all but one of the eicosanoids associated with blood pressure.

The Cox regression models using the eicosanoid risk score replicated in the FHS cohort, but did not quite reach statistical significance in DILGOM. However, a meta-analysis of the risk scores in the three cohorts produced a statistically significant result. These results support our hypothesis of an eicosanoid risk score predicting incident type 2 diabetes. Pathogenetic mechanisms for the effects of these eicosanoids on the risk of incident type 2 diabetes are not known. 12-HHTrE was moderately associated with HOMA-IR and other indicators of insulin resistance, even after adjustment for age, sex and obesity. Interestingly, the eicosanoid 8-iso-PGA1 that replicated strongly in the FHS cohort, did not correlate with glucose levels and only weakly with the insulin indicators.

Our study showed very little correlation between a panel of cytokines and the three eicosanoids significantly associated with type 2 diabetes. In addition, the Cox models further adjusted for cytokines in the FINRISK cohort, remained statistically significant both for the continuous eicosanoid risk score as well as the upper risk quartiles. This suggests that the peptide markers of inflammation, hs-CRP, IL-6, IL-1Ra and TNF-alpha, may not comprehensively reflect all aspects of subclinical inflammation that play a role in risk of type 2 diabetes and can be identified by using eicosanoids as more proximal markers of inflammatory activity. Alternatively, the eicosanoid score may increase the risk of type 2 diabetes through mechanisms other than traditional inflammatory pathways.

Recently it has been suggested that inflammation should be targeted with novel therapeutics, as part of treatment and prevention of type 2 diabetes and its complications, especially because some antidiabetic agents have direct and secondary anti-inflammatory properties [39,40]. Biological and pharmacological characteristics of upstream mediators of inflammation, such as of the two eicosanoids positively associated with incident type 2 diabetes (8-iso-PGA1 and 12-HHTrE), need to be further investigated in order to establish potential eicosanoid-targeting anti-inflammatory therapeutic measures.

Our study suggests potential windows of opportunity for intervention in terms of both prevention and treatment of type 2 diabetes. We speculate that 8-iso-PGA1 and 12-HHTrE could be used as personalized biomarkers in the identification of people at risk of developing type 2 diabetes. Interestingly, the latter metabolite was also associated with hypertension in our recent analysis [36]. Better understanding of determinants and functions of these two eicosanoids may allow for a more targeted approach in the preventive measures such as lifestyle and medical interventions [3,4]. These two eicosanoids, the three-eicosanoid risk score and their novel biomarker potential should be examined with closer detail in order to establish their roles in the early development of insulin resistance and type 2 diabetes.

### Strengths and limitations

Our study is a large population-based random sample of 25 to 74-year-old adults with a high participation rate, prospective design and negligible loss to follow-up. Analyses were controlled for traditional risk factors for incident type 2 diabetes, other factors that may affect inflammation levels and hs-CRP. The partial replication of our results in FHS and DILGOM data sets further strengthens the study. 8-iso-PGA1 replicated robustly in the FHS cohort. The other two were directionally consistent but the lack of statistical significance may be due to the limited statistical power in both FHS and DILGOM. Also, the FHS participants were on average 20 years older than FINRISK participants, which may have had an effect on replication. The eicosanoid profiling for all three cohorts was done in the same laboratory with the same methods. We used a sophisticated, directed, non-targeted LC-MS combined with computational chemical networking to identify hundreds of known and putative eicosanoids from the majority of the study participants.

Our study included mainly white participants of European ancestry and, therefore, may not be generalizable to all ethnic groups. Limitations of our study also include possible bias due to self-reported smoking status and the level of physical activity. Using family history of type 2 diabetes as our only consideration of genetic risk factors instead of e.g. a polygenic risk score, is a further limitation. In addition, we could only detect known and putative eicosanoids that circulate in human plasma. And finally, the identity of several eicosanoids, including one of the eicosanoids with a significant inverse association with our outcome, remains unknown.

### Conclusions

Plasma eicosanoids are associated with incident type 2 diabetes and our three-eicosanoid risk score was an independent predictor of future type 2 diabetes in the general population. Of individual eicosanoids, 8-iso-PGA1 clearly replicated in two independent populations. Furthermore, 12-HHTrE which has been associated with hypertension earlier, predicted type 2 diabetes in the two Finnish cohorts and was close to statistical significance in FHS as well. Finally, our findings provide novel biological information on the development of type 2 diabetes suggesting opportunities for early identification of individuals at risk and potential therapeutic targets for more precise prevention and treatment of type 2 diabetes.

## Supporting information

STROBE checklist

## Data Availability

Data are available from the THL Biobank, based on a written application and relevant Finnish legislation as described on the website of the Biobank https://thl-biobank.elixir-finland.org/.

## ACKNOWLEDGMENTS

We would like to thank the FINRISK 2002 cohort participants as well as participants in the FHS who have contributed to this research for decades.

## CONTRIBUTIONS

KT, JP, VS, MJ, SC, TN, PJ and ASH designed the study. JP and TL performed the statistical analyses and KT drafted the manuscript. JDW, KM, KAL, AA, and MJ performed eicosanoid measurements. All authors contributed to the revision of the manuscript with important intellectual content.

## REFERENCES

[1] GBD 2016 Disease and Injury Incidence and Prevalence Collaborators. Global, regional, and national incidence, prevalence, and years lived with disability for 328 diseases and injuries for 195 countries, 1990-2016: a systematic analysis for the Global Burden of Disease Study 2016 [published correction appears in Lancet. 2017 Oct 28;390(10106):e38]. Lancet. 2017;390(10100):1211–1259. doi:10.1016/S0140-6736(17)32154-2

[2] Bommer C, Sagalova V, Heesemann E, et al. Global Economic Burden of Diabetes in Adults: Projections From 2015 to 2030. Diabetes Care. 2018 May;41(5):963–970. doi: 10.2337/dc17-1962

[3] Tuomilehto J, Lindström J, Eriksson JG, et al. Prevention of type 2 diabetes mellitus by changes in lifestyle among subjects with impaired glucose tolerance. N Engl J Med. 2001;344(18):1343–1350. doi:10.1056/NEJM200105033441801

[4] Knowler WC, Barrett-Connor E, Fowler SE, et al. Reduction in the incidence of type 2 diabetes with lifestyle intervention or metformin. N Engl J Med. 2002;346(6):393–403. doi:10.1056/NEJMoa012512

[5] Festa A, D’Agostino R Jr, Tracy RP, Haffner SM; Insulin Resistance Atherosclerosis Study. Elevated levels of acute-phase proteins and plasminogen activator inhibitor-1 predict the development of type 2 diabetes: the insulin resistance atherosclerosis study. Diabetes. 2002;51(4):1131–1137. doi:10.2337/diabetes.51.4.1131

[6] Herder C, Baumert J, Thorand B, et al. Chemokines as risk factors for type 2 diabetes: results from the MONICA/KORA Augsburg study, 1984-2002. Diabetologia. 2006;49(5):921–929. doi:10.1007/s00125-006-0190-y

[7] Salomaa V, Havulinna A, Saarela O, et al. Thirty-one novel biomarkers as predictors for clinically incident diabetes. PLoS One. 2010;5(4):e10100. Published 2010 Apr 9. doi:10.1371/journal.pone.0010100

[8] Brahimaj A, Ligthart S, Ghanbari M, et al. Novel inflammatory markers for incident pre-diabetes and type 2 diabetes: the Rotterdam Study. Eur J Epidemiol. 2017 Mar;32(3):217–226. doi: 10.1007/s10654-017-0236-0. Epub 2017 Mar 3. PMID: 28258520; PMCID: PMC5380703.

[9] Goldberg RB, Bray GA, Marcovina SM, et al. Non-traditional biomarkers and incident diabetes in the Diabetes Prevention Program: comparative effects of lifestyle and metformin interventions. Diabetologia. 2019;62(1):58–69. doi:10.1007/s00125-018-4748-2

[10] Dennis EA, Norris PC. Eicosanoid storm in infection and inflammation. Nat Rev Immunol. 2015 Aug;15(8):511-23. doi: 10.1038/nri3859. Epub 2015 Jul 3. Erratum in: Nat Rev Immunol. 2015 Nov;15(11):724. PMID: 26139350; PMCID: PMC4606863.

[11] Watrous JD, Niiranen TJ, Lagerborg KA, et al. Directed Non-targeted Mass Spectrometry and Chemical Networking for Discovery of Eicosanoids and Related Oxylipins. Cell Chem Biol. 2019;26(3):433-442.e4. doi:10.1016/j.chembiol.2018.11.015

[12] Lagerborg KA, Watrous JD, Cheng S, Jain M. High-Throughput Measure of Bioactive Lipids Using Non-targeted Mass Spectrometry. Methods Mol Biol. 2019;1862:17–35. doi:10.1007/978-1-4939-8769-6_2

[13] Borodulin K, Tolonen H, Jousilahti P, et al. Cohort Profile: The National FINRISK Study. Int J Epidemiol. 2018;47(3):696–696i. doi:10.1093/ije/dyx239

[14] Kannel WB, Feinleib M, McNamara PM, Garrison RJ, Castelli WP. An investigation of coronary heart disease in families. The Framingham offspring study. Am J Epidemiol. 1979;110:281–290.

[15] Konttinen H, Llewellyn C, Silventoinen K, et al. Genetic predisposition to obesity, restrained eating and changes in body weight: a population-based prospective study. Int J Obes 2018; 42: 858–65. doi: 10.1038/ijo.2017.278

[16] Saaristo T, Peltonen M, Lindström J, et al. Cross-sectional evaluation of the Finnish Diabetes Risk Score: a tool to identify undetected type 2 diabetes, abnormal glucose tolerance and metabolic syndrome. Diab Vasc Dis Res. 2005;2(2):67–72. doi:10.3132/dvdr.2005.011

[17] Matthews DR, Hosker JP, Rudenski AS, Naylor BA, Treacher DF, Turner RC. Homeostasis model assessment: insulin resistance and beta-cell function from fasting plasma glucose and insulin concentrations in man. Diabetologia. 1985;28(7):412–419. doi:10.1007/BF00280883

[18] Sliz E, Kalaoja M, Ahola-Olli A, et al. Genome-wide association study identifies seven novel loci associating with circulating cytokines and cell adhesion molecules in Finns. J Med Genet. 2019;56(9):607–616. doi:10.1136/jmedgenet-2018-105965

[19] Ahola-Olli, AV, Würtz P, Havulinna AS, et al. Genome-wide Association Study Identifies 27 Loci Influencing Concentrations of Circulating Cytokines and Growth Factors. Am J Hum Genet. 2017;100(1):40–50. doi: 10.1016/j.ajhg.2016.11.007

[20] Drotleff B, Lämmerhofer M. Guidelines for Selection of Internal Standard-Based Normalization Strategies in Untargeted Lipidomic Profiling by LC-HR-MS/MS. Anal Chem. 2019;91(15):9836–9843. doi:10.1021/acs.analchem.9b01505

[21] Grambsch PM, Therneau TM. Proportional hazards tests and diagnostics based on weighted residuals, Biometrika, 1994;81(3):515–526. doi:10.1093/biomet/81.3.515.

[22] Glickman ME, Rao SR, Schultz MR. False discovery rate control is a recommended alternative to Bonferroni-type adjustments in health studies. J Clin Epidemiol. 2014;67(8):850–857. doi:10.1016/j.jclinepi.2014.03.012

[23] Etymologia: Bonferroni correction. Emerg Infect Dis. 2015 Feb. doi: 10.3201/eid2102.et2102

[24] Peto R, Peto J. Asymptotically efficient invariant test procedures. J R Stat Soc [A] 1972;135:185–206

[25] Calle MC, Fernandez ML. Inflammation and type 2 diabetes. Diabetes Metab. 2012;38(3):183–191. doi:10.1016/j.diabet.2011.11.006

[26] Brunner EJ, Kivimäki M, Witte DR, et al. Inflammation, insulin resistance, and diabetes--Mendelian randomization using CRP haplotypes points upstream. PLoS Med. 2008;5(8):e155. doi:10.1371/journal.pmed.0050155

[27] Luo P, Wang MH. Eicosanoids, β-cell function, and diabetes. Prostaglandins Other Lipid Mediat. 2011;95(1-4):1–10. doi:10.1016/j.prostaglandins.2011.06.001

[28] Yang Q, Vijayakumar A, Kahn BB. Metabolites as regulators of insulin sensitivity and metabolism. Nat Rev Mol Cell Biol. 2018;19(10):654–672. doi:10.1038/s41580-018-0044-8

[29] Roberts LJ, Morrow JD. Measurement of F(2)-isoprostanes as an index of oxidative stress in vivo. Free Radic Biol Med. 2000;28(4):505–513. doi:10.1016/s0891-5849(99)00264-6

[30] Odegaard AO, Jacobs DR Jr, Sanchez OA, Goff DC Jr, Reiner AP, Gross MD. Oxidative stress, inflammation, endothelial dysfunction and incidence of type 2 diabetes. Cardiovasc Diabetol. 2016;15:51. Published 2016 Mar 24. doi:10.1186/s12933-016-0369-6

[31] Park K, Gross M, Lee DH, et al. Oxidative stress and insulin resistance: the coronary artery risk development in young adults study. Diabetes Care. 2009;32(7):1302–1307. doi:10.2337/dc09-0259

[32] Vincent AM, Russell JW, Low P, Feldman EL. Oxidative stress in the pathogenesis of diabetic neuropathy. Endocr Rev. 2004;25(4):612–628. doi:10.1210/er.2003-0019

[33] Elmarakby AA, Sullivan JC. Relationship between oxidative stress and inflammatory cytokines in diabetic nephropathy. Cardiovasc Ther. 2012;30(1):49–59. doi:10.1111/j.1755-5922.2010.00218.x

[34] Baig S, Shabeer M, Parvaresh Rizi E, et al. Heredity of type 2 diabetes confers increased susceptibility to oxidative stress and inflammation. BMJ Open Diabetes Res Care. 2020;8(1):e000945. doi:10.1136/bmjdrc-2019-000945

[35] Chocholoušková M, Jirásko R, Vrána D, Gatěk J, Melichar B, Holčapek M. Reversed phase UHPLC/ESI-MS determination of oxylipins in human plasma: a case study of female breast cancer. Anal Bioanal Chem. 2019;411(6):1239–1251. doi:10.1007/s00216-018-1556-y

[36] Palmu J, Watrous JD, Mercader K, et al. Eicosanoid Inflammatory Mediators Are Robustly Associated With Blood Pressure in the General Population. J Am Heart Assoc. 2020 Oct 20;9(19):e017598. doi: 10.1161/JAHA.120.017598.

[37] Mezei Z, Kis B, Gecse A, Telegdy G, Abrahám G, Sonkodi S. Platelet eicosanoids and the effect of captopril in blood pressure regulation. Eur J Pharmacol. 1997;340(1):67–73. doi:10.1016/s0014-2999(97)01402-7

[38] Nieman DC, Gillitt ND, Chen GY, Zhang Q, Sakaguchi CA, Stephan EH. Carbohydrate intake attenuates post-exercise plasma levels of cytochrome P450-generated oxylipins. PLoS One. 2019;14(3):e0213676. Published 2019 Mar 18. doi:10.1371/journal.pone.0213676

[39] Goldfine AB, Shoelson SE. Therapeutic approaches targeting inflammation for diabetes and associated cardiovascular risk. J Clin Invest. 2017;127(1):83–93. doi:10.1172/JCI88884

[40] Donath MY. Multiple benefits of targeting inflammation in the treatment of type 2 diabetes. Diabetologia. 2016;59(4):679–682. doi:10.1007/s00125-016-3873-z

