## Supplemental figures and tables for "A plasma metabolite score of three eicosanoids predicts incident type 2 diabetes – a prospective study in three independent cohorts"

**Supplement Figure S1. Flowcharts representing the exclusion criteria and final study population in discovery analyses and replication cohorts.**

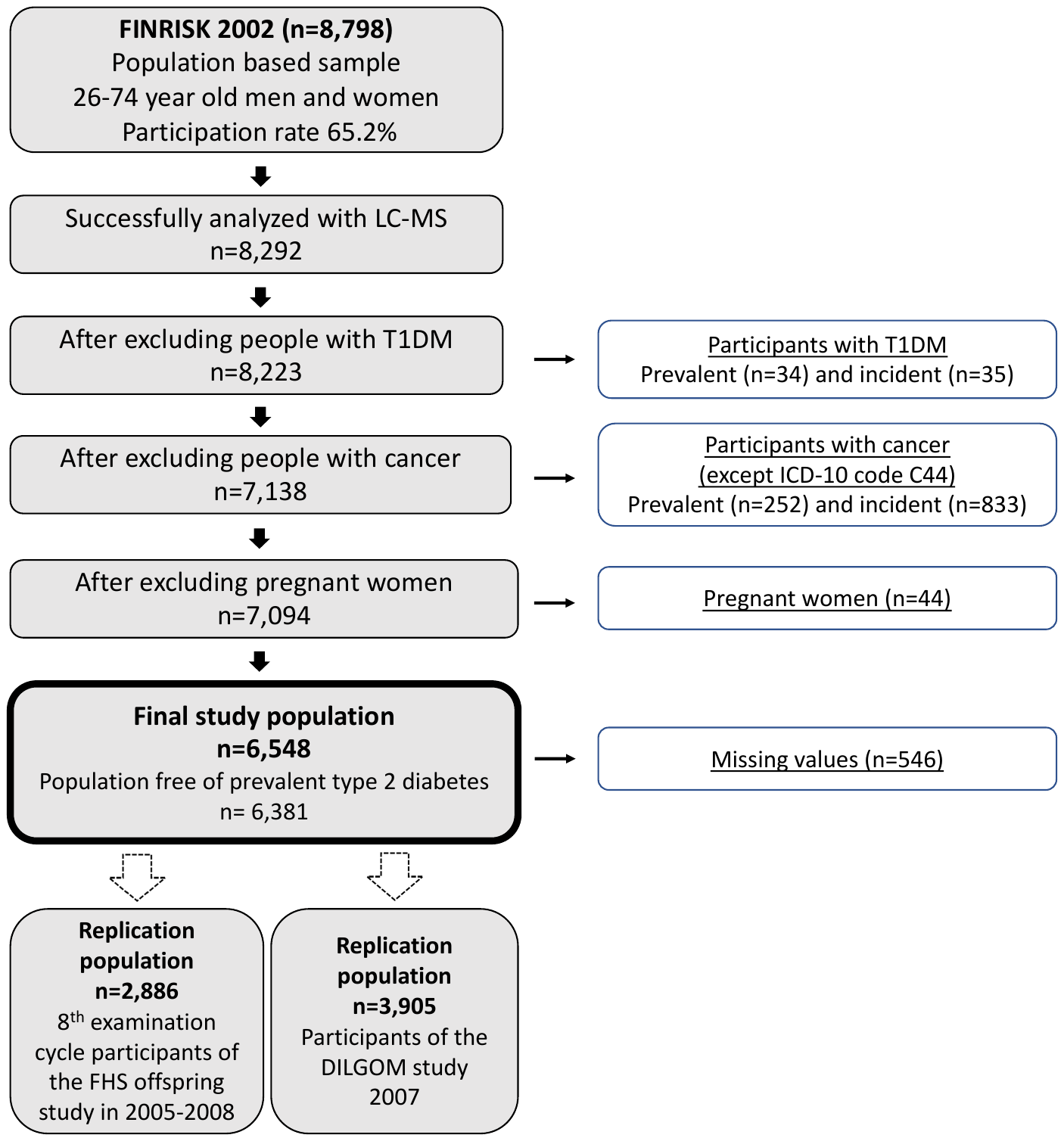

**Supplement Figure S2. Correlation heatmap for the 76 eicosanoids associated with incident type 2 diabetes and hs-CRP**

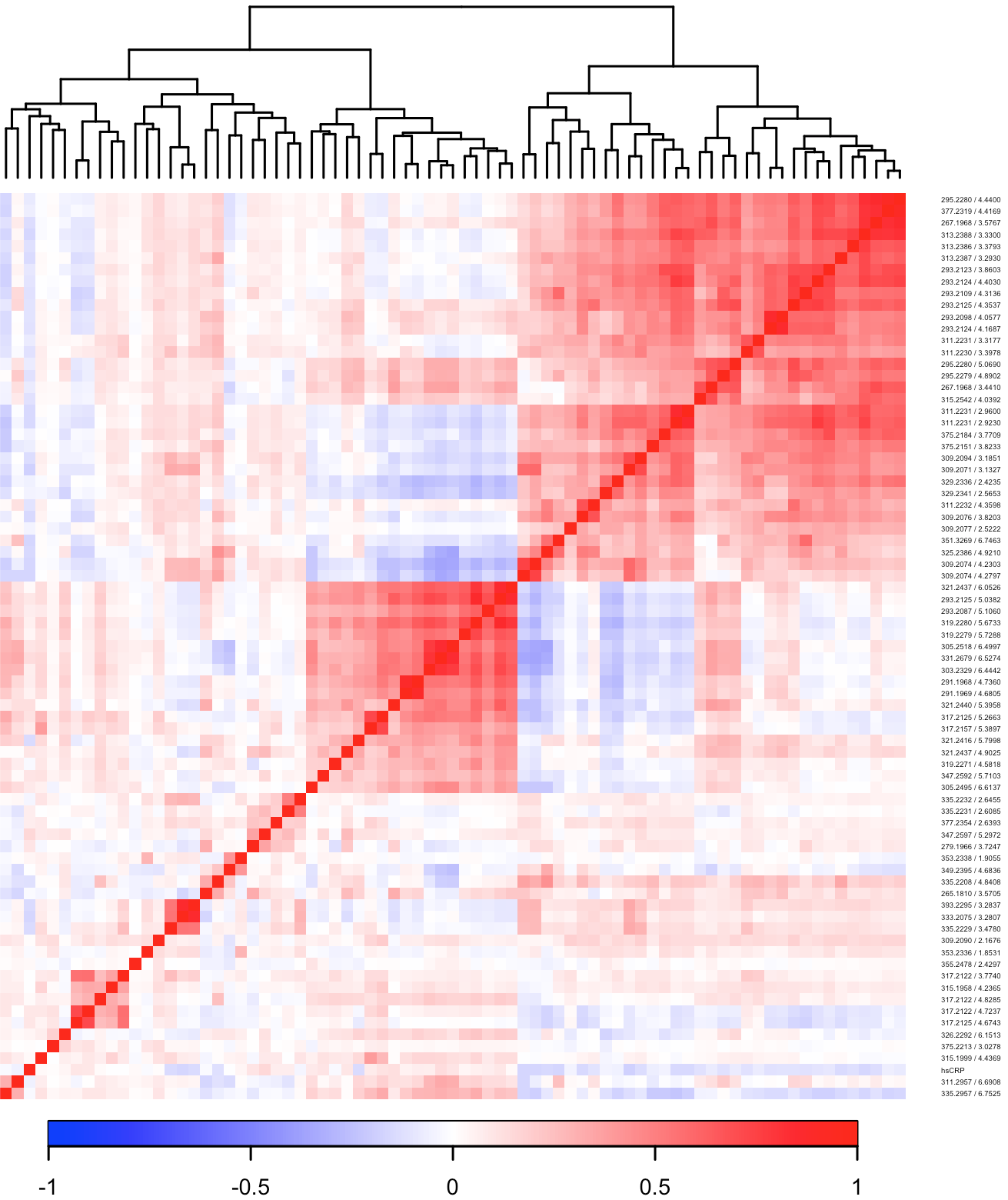

Links between eicosanoids significantly associated with incident T2D, and hs-CRP, were determined using Spearman rank correlation and ordered using the complete linkage clustering method.

**Supplement Table S1**. Medians of fP-Gluc, fP-Insulin, HOMA-IR and HOMA-B in eicosanoid risk score quartiles and test of linear trend across the quartiles in FINRISK 2002.

|  | 1^st^ risk quartile | 2^nd^ risk quartile | 3^rd^ risk quartile | 4^th^ risk quartile | p-value for trend* |
| --- | --- | --- | --- | --- | --- |
| **fP-Gluc, mmol/l** | 5.55 | 5.66 | 5.77 | 5.82 | <0.0001 |
| **fP-Insulin, mmol/l** | 6.5 | 6.5 | 7.5 | 8.7 | <0.0001 |
| **HbA1c, mmol/mol** | 36 | 36 | 36 | 37 | <0.0001 |
| **HOMA-IR** | 1.61 | 1.60 | 1.87 | 2.26 | <0.0001 |
| **HOMA-B** | 63.2 | 61.3 | 65.9 | 74.9 | 0.0140 |
| **hs-CRP, mg/l** | 0.8 | 0.94 | 1.15 | 1.28 | 0.0008 |

Adjusted for age, sex and BMI

**Supplement Table S2**. Spearman rank correlations of the three eicosanoids used for the risk score with glucose tolerance indicators and inflammation markers in FINRISK 2002.

|  | **311.223100_2.9230**  **Unknown eicosanoid** | **335.223200_2.6455**  **8-iso-PGA1** | **279.196600_3.7247**  **12-HHTrE** |
| --- | --- | --- | --- |
| **fP-Gluc** | -0.11* | 0.03 | 0.15* |
| **fP-Insulin** | -0.07 | 0.10* | 0.24* |
| **HbA1c** | -0.08 | 0.07 | 0.11* |
| **HOMA-IR** | -0.08 | 0.10* | 0.25* |
| **HOMA-B** | -0.02 | 0.09* | 0.18* |
| **hs-CRP** | -0.14* | -0.02 | -0.09* |
| **IL-1RA** | 0.00 | -0.02 | -0.01 |
| **IL-6** | 0.01 | 0.03 | 0.06 |
| **TNF-alpha** | 0.02 | -0.01 | 0.01 |

*p<0.0001
