## Supplementary material for "A plasma metabolite score of three eicosanoids predicts incident type 2 diabetes – a prospective study in three independent cohorts": STROBE checklist

STROBE Statement—Checklist of items that should be included in reports of ***cohort studies***

|  | **Item No** | **Recommendation** | **Reference in manuscript** |
| --- | --- | --- | --- |
| **Title and abstract** | 1 | (*a*) Indicate the study’s design with a commonly used term in the title or the abstract (*b*) Provide in the abstract an informative and balanced summary of what was done and what was found | a) Title: […] prospective study in three independent cohorts  b) Methods and Results subsections of the abstract: eg. […] In the FINRISK 2002 study, a population-based sample of Finnish men and women aged 25-74 years, we used directed, non-targeted liquid chromatography – mass spectrometry to identify 545 eicosanoids and related oxylipins in the participants’ plasma samples (n=8,292). We used multivariable-adjusted Cox regression to examine associations between eicosanoids and incident type 2 diabetes. The findings were replicated in the Framingham Heart Study (FHS, n=2,886) and DILGOM 2007 (n=3,905). Together, these three cohorts had 1070 cases of incident type 2 diabetes. […] |
| **Introduction** | | | |
| Background/  rationale | 2 | Explain the scientific background and rationale for the investigation being reported | Introduction: E.g. Type 2 diabetes is closely linked to […], all of which are known to be associated with low-grade inflammation. Biomarkers of systemic inflammation […] associated with the development of type 2 diabetes in large population-based studies and randomized controlled trials. […] Eicosanoids mediate inflammatory processes directly at local level as well as systemically […]. With recent advances in mass spectrometry-based metabolomics, hundreds of eicosanoid and oxylipin species can now be detected and quantified at a rapid scale in human plasma […] |
| Objectives | 3 | State specific objectives, including any prespecified hypotheses | Introduction: E.g. The aim of this study was to examine whether plasma eicosanoid profiles are associated with risk of incident type 2 diabetes in a prospective follow-up of a population-based Finnish cohort. […] A secondary aim was to investigate whether the eicosanoid effect is mediated by inflammation or insulin resistance. |
| **Methods** | | | |
| Study design | 4 | Present key elements of study design early in the paper | Material and Methods: E.g. […] prospective follow-up of a population-based Finnish cohort. The significant findings were replicated in an independent cohort from the North American sub-continent and another independent population-based Finnish cohort. |
| Setting | 5 | Describe the setting, locations, and relevant dates, including periods of recruitment, exposure, follow-up, and data collection | Material and Methods: E.g. For the FINRISK 2002 cohort, prevalent diabetes cases at baseline and incident cases during follow-up were identified combining information from the National Hospital Discharge Register (NHDR), Causes of Death Register (CDR) and the Drug Reimbursement and Drug Purchase Registers (DPR), using the Finnish personal identification number, up to December 31^st^, 2017. […] |
| Participants | 6 | (*a*) Give the eligibility criteria, and the sources and methods of selection of participants. Describe methods of follow-up; | a) Material and Methods: E.g. […] FINRISK 2002 study is a population-based random sample of individuals aged 25-74 years living in Finland (n=8798, participation rate 65.2%). […] Participants with prevalent or incident type 1 diabetes were excluded from the analysis (n=79). Record linkages based on the personal identification code to NHDR, CDR and DPR were also used to identify subjects to be excluded from the analysis, due to cancer (excluding ICD10 category C44, n=1,085). […] |
| Variables | 7 | Clearly define all outcomes, exposures, predictors, potential confounders, and effect modifiers. Give diagnostic criteria, if applicable | Material and Methods, Results table 1, etc: E.g. For the FINRISK 2002 [and the DILGOM 2007] Cohort[s] […] to determine the type of diabetes, we used a proxy variable: all participants under 30 years of age and treated with insulin only, or in combination with metformin, and those aged 30-40 years when insulin only was started, were categorized as type 1 diabetes. All other persons with diabetes were considered to have type 2 diabetes. […] Diabetes mellitus in FHS Offspring cohort was diagnosed either by fasting plasma glucose ≥126 mg/dL, non-fasting plasma glucose ≥200 mg/dL, or treatment with insulin or an oral hypoglycemic agent as ascertained at routine FHS examinations or based on annual medical health history updates. |
| Data sources /measurement | 8 | For each variable of interest, give sources of data and details of methods of assessment (measurement). Describe comparability of assessment methods if there is more than one group | Material and Methods: E.g. […] The OGTT was carried out according to the World Health Organization (WHO) recommendations, and the testing and measurement methodologies have previously been described in detail [15]. Fasting plasma glucose and insulin concentrations were used to calculate the homeostasis model assessment for insulin resistance (HOMA-IR) and for beta cell function (HOMA-B) indices [16]. |
| Bias | 9 | Describe any efforts to address potential sources of bias | Material and Methods etc: E.g. Participants […] underwent physical examination by trained nurses […] In total, 545 eicosanoids and related oxylipins were validated using methodologies such as spectral fragmentation pattern networking and manual annotation. […] |
| Study size | 10 | Explain how the study size was arrived at | Material and Methods, Supplement figure 1 (flowchart of participants): E.g. […] Supplement Figure S1 presents a flowchart formulating the final study sample. |
| Quantitative variables | 11 | Explain how quantitative variables were handled in the analyses. If applicable, describe which groupings were chosen and why | Material and Methods: E.g. The eicosanoid profiling data was normalized using plate medians corrected for plate deviation: plate medians were subtracted from each feature and then divided by the median absolute deviation […] we used means, or where relevant due to skewed distributions, geometrical means, and interquartile ranges to summarize baseline characteristics of continuous variables and frequencies for categorical variables. […] |
| Statistical methods | 12 | *a*) Describe all statistical methods, including those used to control for confounding; *b*) Describe any methods used to examine subgroups and interactions; *c*) Explain how missing data were addressed; *d*) If applicable, explain how loss to follow-up was addressed; *e*) Describe any sensitivity analyses | a) Material and Methods, Statistical methods subsection: E.g. […] To test the association of eicosanoids with incident type 2 diabetes, we used Cox proportional hazards regression and a nested modeling approach, adjusting for well-established risk factors for type 2 diabetes, and other confounding factors, in three different models. […]; b) Material and Methods, Table 1 (men and women) etc: E.g. […] Sampling included stratification by sex, region and 10-year age groups. […]; c) Material and Methods Statistical methods subsection: E.g. […] Finally, after excluding further participants with missing values of variables relevant for our analyses […] Participants with baseline diabetes (n=302) and missing values in the covariates were excluded from the analysis, resulting in n=236 incident diabetes cases out of a total of 2,115 individuals included. […] the source code for analyses is available at <https://doi.org/10.5281/zenodo.3968712>.; d) NA; e) NA |
| **Results** | | | |
| Participants | 13 | (a) Report numbers of individuals at each stage of study—eg numbers potentially eligible, examined for eligibility, confirmed eligible, included in the study, completing follow-up, and analysed; b) Give reasons for non-participation at each stage; c) Consider use of a flow diagram | a+b+c) Material and Methods, Table 1, Supplement figure 1 (flowchart of participants) etc: E.g. Supplement Fig S1 presents a flowchart formulating the final study sample. […] HbA1c values were only available for FINRISK participants older than 50 years (n=3,586). […] |
| Descriptive data | 14 | (a) Give characteristics of study participants (eg demographic, clinical, social) and information on exposures and potential confounders; b) Indicate number of participants with missing data for each variable of interest; c) Summarise follow-up time (eg, average and total amount) | a+b) Results, Table 1; c) Results, Figure 3; E.g. […] For the FINRISK 2002 cohort, prevalent diabetes cases at baseline and incident cases during follow-up were identified […] using the Finnish personal identification number, up to December 31^st^, 2017. […] Kaplan-Meier curves for the risk score quartiles in FINRISK are presented in Figure 3. |
| Outcome data | 15 | Report numbers of outcome events or summary measures over time | Abstract, Introduction, Material and Methods, Table 1: E.g. […] Together, these cohorts included 1070 cases of incident type 2 diabetes. |
| Main results | 16 | a) Give unadjusted estimates and, if applicable, confounder-adjusted estimates and their precision (eg, 95% confidence interval). Make clear which confounders were adjusted for and why they were included; b) Report category boundaries when continuous variables were categorized; c) If relevant, consider translating estimates of relative risk into absolute risk for a meaningful time period | a) Results, Figures 1, 2, 3, 4: E.g. […]The multivariate adjusted hazard ratio per SD for the three different cohorts are presented in Fig 3, together with the results of a random effects meta-analysis of the risk scores in the three cohorts […]; b+c) NA |
| Other analyses | 17 | Report other analyses done—eg analyses of subgroups and interactions, and sensitivity analyses | Last subsection of results: e.g. None of the three eicosanoids correlated with the blood concentrations of inflammatory cytokines. In addition, Cox proportional hazards regression analysis for the multivariable model, further adjusted for the inflammatory cytokines, remained significant for both the continuous three-eicosanoid risk score as well as the 3rd and 4th risk quartiles of the score. |
| **Discussion** | | | |
| Key results | 18 | Summarise key results with reference to study objectives | Discussion: e.g. Combined with advanced metabolomics methodologies, our statistically cogent survival analysis using a three-eicosanoid risk score suggests a significant independent role for lipid-derived, upstream mediators of inflammation in the prediction of incident type 2 diabetes. […] |
| Limitations | 19 | Discuss limitations of the study, taking into account sources of potential bias or imprecision. Discuss both direction and magnitude of any potential bias | Discussion: e.g. […] Limitations of our study also include possible bias due to self-reported smoking status and the level of physical activity. Using family history of type 2 diabetes as our only consideration of genetic risk factors instead of e.g. a polygenic risk score, is a further limitation. In addition, we could only detect known and putative eicosanoids that circulate in human plasma. […] |
| Interpretation | 20 | Give a cautious overall interpretation of results considering objectives, limitations, multiplicity of analyses, results from similar studies, and other relevant evidence | Discussion, and conclusions subsection: e.g. […] Finally, our findings provide opportunities for early identification of individuals at risk of type 2 diabetes and potential therapeutic targets for prevention and treatment of type 2 diabetes. |
| Generalisa-bility | 21 | Discuss the generalisability (external validity) of the study results | Discussion: e.g. Our study included mainly white participants of European ancestry and, therefore, may not be generalizable to all ethnic groups. |
| **Other information** | | | |
| Funding | 22 | Give the source of funding and the role of the funders for the present study and, if applicable, for the original study on which the present article is based | Funding section: e.g. […] This work was supported by Paavo Nurmi, Aarne Koskelo, Emil Aaltonen and Finnish Medical Foundations. […] etc |
